# Heterogeneous effects of antidepressants on suicidal ideation: role of polygenic scores

**DOI:** 10.1101/2024.11.02.24316657

**Authors:** Ryunosuke Goto, Tatsuhiko Naito, Norbert Skokauskas, Kosuke Inoue

**Affiliations:** Department of Biomedical Data Science, Stanford University, Stanford, CA, USA; Nash Family Department of Neuroscience & Friedman Brain Institute, Icahn School of Medicine at Mount Sinai, New York, NY, USA; Department of Genetics and Genomic Sciences, Icahn School of Medicine at Mount Sinai, New York, NY, USA; Icahn Institute for Data Science and Genomic Technology, Icahn School of Medicine at Mount Sinai, New York, NY, USA; Ronald M. Loeb Center for Alzheimer’s Disease, Icahn School of Medicine at Mount Sinai, New York, NY, USA; New York Genome Center, New York, NY, USA; Regional Centre for Children and Youth Mental Health and Child Welfare - Central Norway, IPH, Norwegian University of Science and Technology, Trondheim, Norway; Department of Health Promotion and Human Behavior, Graduate School of Medicine, Kyoto University, Kyoto, Japan

## Abstract

Depression continues to be a major contributor to the global burden of diseases. Antidepressants are recommended as the initial choice of treatment for moderate and severe depression among adults, but the choice of antidepressant class can be challenging, as efficacies of commonly used antidepressants are generally comparable between classes. As such, to determine which medication to prescribe to each patient, clinicians often rely on the adverse effect profile of antidepressants. One of the most discussed adverse effects is the possibility of increased suicidality with antidepressant initiation, a rare but serious adverse event. Efforts to predict this have mostly relied on clinical and sociodemographic characteristics of patients, but we do not yet have a clear picture. A promising avenue is to use patients’ genetic data, but the evidence on its utility has so far been mixed, and few large scale databases have both clinical and genetic data that can evaluate its utility. Here, using genetic and clinical data on more than 7,000 patients, both children and adults, with major depressive disorder from the All of Us Research Program, we show that the genetic predisposition for psychiatric disorders may underlie the substantial heterogeneity in the risk of suicidal thoughts with antidepressant use. Specifically, using a target trial emulation framework, we show that patients with higher polygenic scores for psychiatric disorders, particularly for attention deficit-hyperactivity disorder, may be more likely than those with lower scores to experience suicidal thoughts with the use of selective serotonin reuptake inhibitors, relative to bupropion. In the personalized medicine framework, PGSs for various psychiatric disorders may help tailor antidepressants to each patient to avoid serious adverse effects such as suicidal thoughts.

## Main

Depression is a psychiatric disorder that continues to put large burden worldwide [1]. Several evidence-based treatments exist for the disorder, including pharmacotherapy, psychotherapy, electroconvulsive therapy, and transcranial magnetic stimulation. Antidepressants are recommended as the initial choice of treatment for moderate and severe depression among adults. Because the efficacies of commonly used antidepressants are generally comparable between classes, the choice of antidepressants will largely be based on the adverse effect profile [2]. However, the challenge lies in predicting the adverse effects, which is essential in determining which patients to prescribe the most commonly-used selective serotonin reuptake inhibitors (SSRIs) or to prescribe alternative medications such as serotonin and norepinephrine reuptake inhibitors (SNRIs) or bupropion. This is a question that has come up repeatedly in clinical psychiatry, but one without a clear answer [3]. Currently, finding the appropriate medication for each patient usually requires trial-and-error, and the delayed therapeutic effects of antidepressants makes this even more challenging [4].

Among the adverse effect profiles of antidepressants, perhaps one that has received the most attention is the possibility of increased suicidality with antidepressant initiation, the so called “black-box” warning of antidepressants by the Food and Drug Administration [5, 6]. As a result, clinicians have sought to predict suicidal behavior among antidepressant users, first using patients’ clinical and sociodemographic characteristics. Though several potential predictors have been found, conclusive evidence is still lacking [3, 7]. One exception may be younger age, which has been reported to be associated with increased suicidal behavior with antidepressant initiation [8], but this has sparked considerable debate, with some calls to remove the “black-box” warning amidst declining antidepressant prescription rates that led to the Food and Drug Administration’s modification of the warning [5]. As such, better predictors of antidepressants’ adverse events are needed to better inform practice.

Researchers have thus turned to patients’ genetic information, including polygenic scores (PGSs), to better understand factors that may help predict antidepressant response. This may be a promising avenue given the growing evidence that psychiatric disorders, including depression, are typically complex traits, with variants associated with these diseases dispersed across the allelic spectrum [9]. There have been many studies on the link between the effectiveness of antidepressants and PGSs for depression. For instance, studies have pointed to associations between a higher PGS for major depressive disorder and a prescription of more than two antidepressants [10] as well as changes in depression scores with SSRI use [11].

There have also been attempts to guide antidepressant choice with PGSs for other psychiatric disorders, namely schizophrenia, bipolar disorder, and attention-deficit hyperactivity disorder (ADHD), each of which shares genetic factors with depression [12, 13]. For instance, PGS for schizophrenia was not associated with changes in depression scores after treatment with SSRIs, though the study may have been underpowered [14]. Though some suggest that non-response to antidepressants may be due to undiagnosed bipolar disorder, bipolar disorder PGS was not associated with antidepressant response [15]. Finally, studies on PGSs for ADHD, a common comorbidity of major depressive disorder [16], was associated with higher odds of prescription of multiple antidepressants [10].

These findings, though valuable, have mostly focused on antidepressant response and less on the adverse effects of antidepressants. However, clinicians often base their choice of antidepressants on the possibility of serious adverse events such as suicidal behavior, especially when prescribing to young people [5]. As such, we aimed to provide evidence on the effects of different types of antidepressants with the goal of advancing precision/genomic medicine and guiding clinical practice. We focus on antidepressants that are primarily used in clinical practice: SSRIs, SNRIs, and bupropion [2]. Importantly, SNRIs and bupropion are viable alternatives to SSRIs, which are often considered to be first-line antidepressants; thus, this study can inform for whom these medications should be used in place of SSRIs. Equally as important, we evaluate the antidepressants’ effects on suicidal ideation, which is of utmost clinical importance both as a primary outcome (as one of the main symptoms of depression) and as a potential adverse event.

## Results

### Target trial emulation of antidepressants

We used the target trial emulation framework [17] on All of Us Research Program data [18] to compare the effects of three types of antidepressants (SSRIs, SNRIs, and bupropion) on suicidal thoughts, across demographic and clinical characteristics as well as PGSs for psychiatric disorders. A total of 7,848 participants were matched for SSRIs vs. bupropion and 9,316 participants were matched for SSRIs vs. SNRIs, and both samples included children and adults. In the SSRIs vs. bupropion sample, 68.0% were female, the mean age was 47.8 years (standard deviation (SD), 14.4), and 11.3% identified as Hispanic and 14.4% as Non-Hispanic Black. Among the previous diagnoses, anxiety was the most prevalent at 9.4%. In the SSRIs vs. SNRIs sample, 71.1% were female, the mean age was 49.6 (standard deviation (SD), 14.3), and 12.2% identified as Hispanic and 15.1% as Non-Hispanic Black. Among the previous diagnoses surveyed, anxiety was the most prevalent at 9.0%. Both the SSRIs vs. bupropion and SSRIs vs. SNRIs samples achieved good balance of covariate distributions, including previous diagnoses of psychiatric disorders and PGSs (Tables 1 and 2).

**Table 1:** **Comparison of matched bupropion and SSRI samples*^a^***

|  | Bupropion<br>n = 3924 | SSRI<br>n = 3924 | SMD |
| --- | --- | --- | --- |
| Gender (%) |  |  | 0.048 |
| Female | 2631 (67.0) | 2704 (68.9) |  |
| Male | 1267 (32.3) | 1187 (30.2) |  |
| Other | 26 ( 0.7) | 33 ( 0.8) |  |
| Age (mean (SD)) | 47.93 (14.08) | 47.58 (14.76) | 0.024 |
| Race and ethnicity (%) |  |  | 0.099 |
| Hispanic | 391 (10.0) | 497 (12.7) |  |
| Non-Hispanic Black | 542 (13.8) | 590 (15.0) |  |
| Non-Hispanic White | 2814 (71.7) | 2659 (67.8) |  |
| Other | 177 ( 4.5) | 178 ( 4.5) |  |
| College education or higher (%) | 3030 (77.2) | 2906 (74.1) | 0.074 |
| Annual income (%) |  |  | 0.077 |
| -\$25,000 | 1315 (33.5) | 1420 (36.2) | |
| \$25,000-\$50,000 | 785 (20.0) | 830 (21.2) | |
| \$50,000-\$100,000 | 987 (25.2) | 909 (23.2) | |
| \$100,000- | 837 (21.3) | 765 (19.5) | |
| Has health insurance (%) | 3823 (97.4) | 3765 (95.9) | 0.083 |
| Born in the US (%) | 3621 (92.3) | 3555 (90.6) | 0.060 |
| Ever-smoker (%) | 2180 (55.6) | 1975 (50.3) | 0.105 |
| Previous diagnosis of suicidal thoughts (%) | 148 (3.8) | 210 (5.4) | 0.076 |
| Previous diagnosis of anxiety (%) | 338 (8.6) | 396 (10.1) | 0.051 |
| Previous diagnosis of alcohol abuse (%) | 218 (5.6) | 227 (5.8) | 0.010 |
| Previous diagnosis of substance abuse (%) | 183 (4.7) | 198 (5.0) | 0.018 |
| Previous diagnosis of schizophrenia (%) | 54 (1.4) | 52 (1.3) | 0.004 |
| Previous diagnosis of eating disorders (%) | 12 (0.3) | 12 (0.3) | <0.001 |
| Previous diagnosis of PTSD (%) | 244 (6.2) | 254 (6.5) | 0.010 |
| Previous diagnosis of ADHD (%) | 194 (4.9) | 138 (3.5) | 0.071 |
| Previous tricyclic antidepressant use (%) | 351 (8.9) | 339 (8.6) | 0.011 |
| PGS for depression (mean (SD)) | 0.00 (1.03) | 0.02 (0.98) | 0.018 |
| PGS for self-injurious behavior (mean (SD)) | 0.05 (0.99) | 0.05 (1.01) | 0.001 |
| PGS for anxiety (mean (SD)) | -0.01 (1.01) | -0.01 (1.01) | 0.005 |
| PGS for bipolar disorder (mean (SD)) | -0.19 (0.98) | -0.16 (0.98) | 0.029 |
| PGS for schizophrenia (mean (SD)) | -0.29 (0.92) | -0.23 (0.94) | 0.067 |
| PGS for ADHD (mean (SD)) | -0.14 (0.96) | -0.10 (0.97) | 0.037 |

<sup>a</sup> Each PGS was standardized across all All of Us Research Program participants with non-missing race and ethnicity and trait labels prior to analyses. SSRI, serotonin reuptake inhibitor; PTSD, post-traumatic stress disorder; ADHD, attention deficit-hyperactivity disorder; PGS, polygenic score.

**Table 2:** **Comparison of matched SNRI and SSRI samples*^b^***

|  | SNRI<br>n = 4658 | SSRI<br>n = 4658 | SMD |
| --- | --- | --- | --- |
| Gender (%) |  |  | 0.063 |
| Female | 3376 (72.5) | 3248 (69.7) |  |
| Male | 1240 (26.6) | 1371 (29.4) |  |
| Other | 42 ( 0.9) | 39 ( 0.8) |  |
| Age (mean (SD)) | 50.47 (13.80) | 48.69 (14.70) | 0.125 |
| Race and ethnicity (%) |  |  | 0.070 |
| Hispanic | 528 (11.3) | 608 (13.1) |  |
| Non-Hispanic Black | 683 (14.7) | 725 (15.6) |  |
| Non-Hispanic White | 3248 (69.7) | 3104 (66.6) |  |
| Other | 199 ( 4.3) | 221 ( 4.7) |  |
| College education or higher (%) | 3515 (75.5) | 3404 (73.1) | 0.055 |
| Annual income (%) |  |  | 0.048 |
| -\$25,000 | 1885 (40.5) | 1877 (40.3) | |
| \$25,000-\$50,000 | 991 (21.3) | 927 (19.9) | |
| \$50,000-\$100,000 | 1018 (21.9) | 1019 (21.9) | |
| \$100,000- | 764 (16.4) | 835 (17.9) | |
| Has health insurance (%) | 4526 (97.2) | 4490 (96.4) | 0.044 |
| Born in the US (%) | 4288 (92.1) | 4236 (90.9) | 0.040 |
| Ever-smoker (%) | 2380 (51.1) | 2286 (49.1) | 0.040 |
| Previous diagnosis of suicidal thoughts (%) | 208 (4.5) | 271 (5.8) | 0.061 |
| Previous diagnosis of anxiety (%) | 384 (8.2) | 459 (9.9) | 0.056 |
| Previous diagnosis of alcohol abuse (%) | 233 (5.0) | 285 (6.1) | 0.049 |
| Previous diagnosis of substance abuse (%) | 189 (4.1) | 236 (5.1) | 0.048 |
| Previous diagnosis of schizophrenia (%) | 53 (1.1) | 71 (1.5) | 0.034 |
| Previous diagnosis of eating disorders (%) | 7 (0.2) | 13 (0.3) | 0.028 |
| Previous diagnosis of PTSD (%) | 340 (7.3) | 353 (7.6) | 0.011 |
| Previous diagnosis of ADHD (%) | 140 (3.0) | 172 (3.7) | 0.038 |
| Previous tricyclic antidepressant use (%) | 572 (12.3) | 399 (8.6) | 0.122 |
| PGS for depression (mean (SD)) | 0.02 (1.04) | 0.03 (1.00) | 0.015 |
| PGS for self-injurious behavior (mean (SD)) | 0.06 (1.02) | 0.05 (1.01) | 0.016 |
| PGS for anxiety (mean (SD)) | 0.00 (1.02) | 0.00 (1.01) | 0.004 |
| PGS for bipolar disorder (mean (SD)) | -0.18 (0.99) | -0.15 (0.99) | 0.026 |
| PGS for schizophrenia (mean (SD)) | -0.26 (0.93) | -0.21 (0.95) | 0.050 |
| PGS for ADHD (mean (SD)) | -0.11 (0.96) | -0.08 (0.97) | 0.033 |

<sup>b</sup> Each PGS was standardized across all All of Us Research Program participants with non-missing race and ethnicity and trait labels prior to analyses. SNRI, serotonin and norepinephrine reuptake inhibitor; SD, standard deviation; SSRI, serotonin reuptake inhibitor; SD, standard deviation; PTSD, post-traumatic stress disorder; ADHD, attention deficit-hyperactivity disorder; PGS, polygenic score.

The sources of the selected PGSs and links in the PGS catalog are available in Supplementary Table 1. In general, the PGSs achieved modest performance in predicting the psychiatric disorders (AUROCs up to 0.67), though in some cases the AUROCs were close to 0.5. They tended to perform worse in groups other than non-Hispanic Whites (Extended Data Figures 1–6).

We fit Cox proportional hazard models to estimate hazard ratios of the risks of suicidal thoughts associated with antidepresssant use. SSRIs had a higher risk of suicidal thoughts relative to bupropion (hazard ratio, 1.36; 95% confidence interval (CI), 1.14 to 1.63; P-value < 0.001; Figure 1a). In contrast, SSRIs and SNRIs had comparable risks of suicidal thoughts (hazard ratio of SSRIs relative to SNRIs, 1.05; 95%CI, 0.91 to 1.22; P-value = 0.50; Figure 1b)

**Figure 1:**
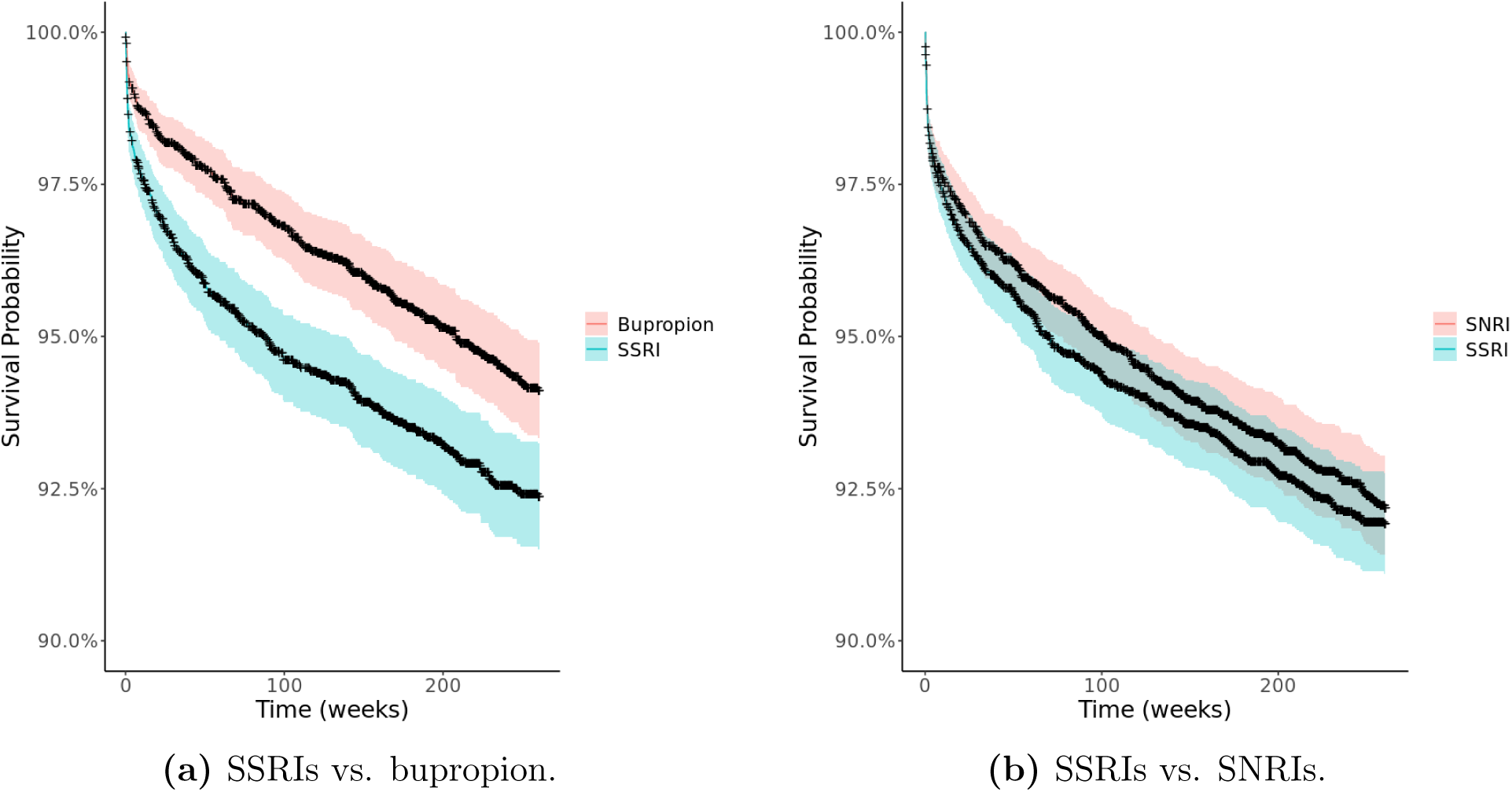
Kaplan-Meier curves of suicidal thoughts.

Subgroup analyses reveal effect heterogeneity across ADHD PGS

We conducted subgroup analyses to evaluate the heterogeneity in the risks of suicidal thoughts associated with antidepressant use across demographic characteristics, clinical characteristics, and PGSs. In the comparison bewteen SSRIs and bupropion, participants generally had similar hazard ratios across nongenetic characteristics (Figure 2). Participants with PGS for ADHD higher than the median had a hazard ratio of 1.66 (95%CI, 1.30 to 2.12), while participants with PGS for ADHD lower than the median had a hazard ratio of 1.06 (95%CI, 0.81 to 1.38; P-for-interaction, 0.01). We observed a similar trend for schizophrenia: participants with PGS for schizophrenia higher than the median had a hazard ratio of 1.52 (95%CI, 1.19 to 1.93), while participants with PGS for schizophrenia lower than the median had a hazard ratio of 1.17 (95%CI, 0.89 to 1.53; P-for-interaction, 0.16; Figure 2).

**Figure 2:**
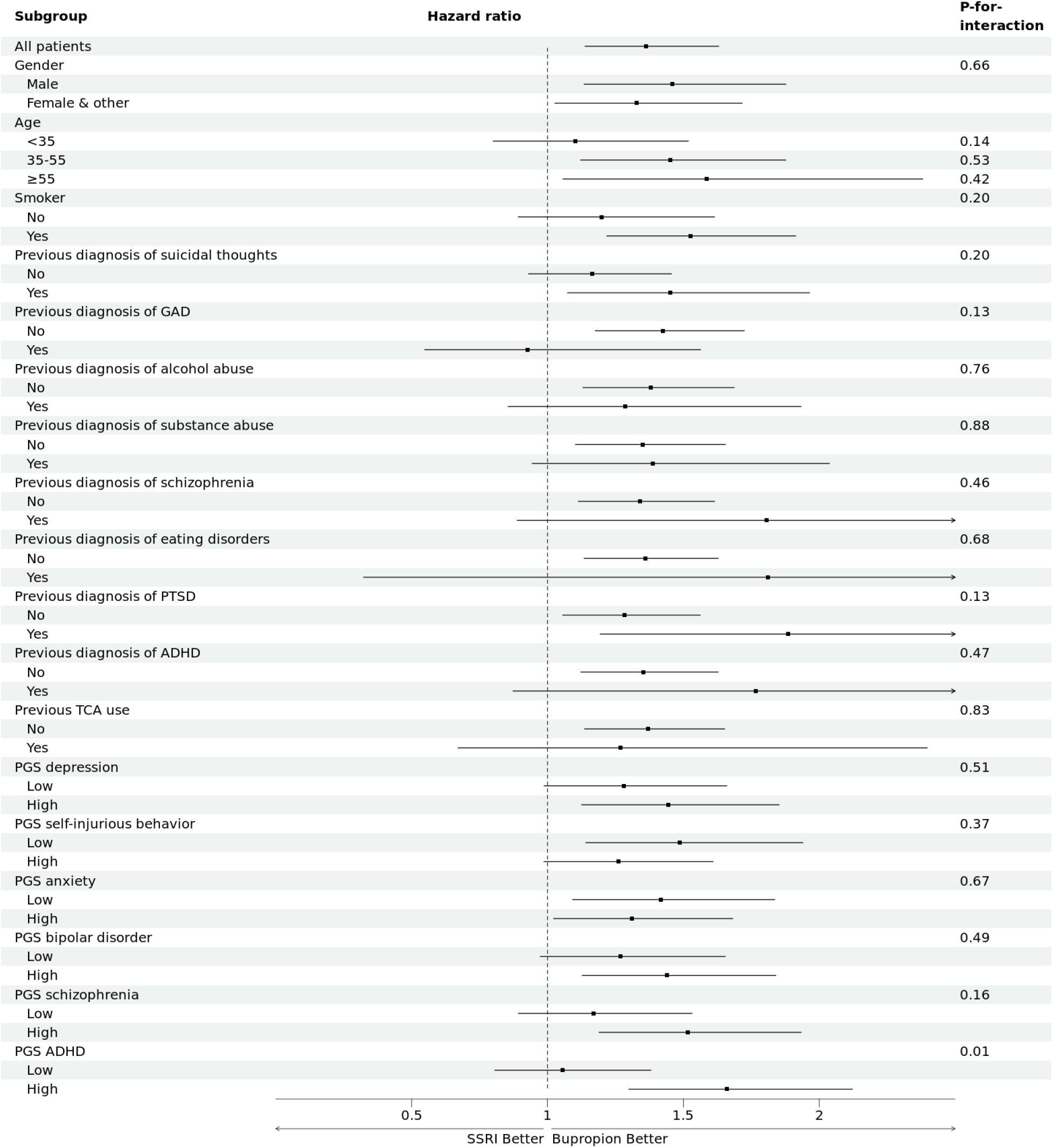
Subgroup analyses for SSRIs vs. bupropion.

In the comparison between SSRIs and SNRIs, males had a hazard ratio of 1.31 (95%CI, 1.04 to 1.65), while other genders had a hazard ratio of 0.86 (95%CI, 0.70 to 1.05; P-for-interaction, 0.01). Hazard ratios were similar across PGSs, with all PGS subgroups having treatment effects close to the null (Figure 3).

**Figure 3:**
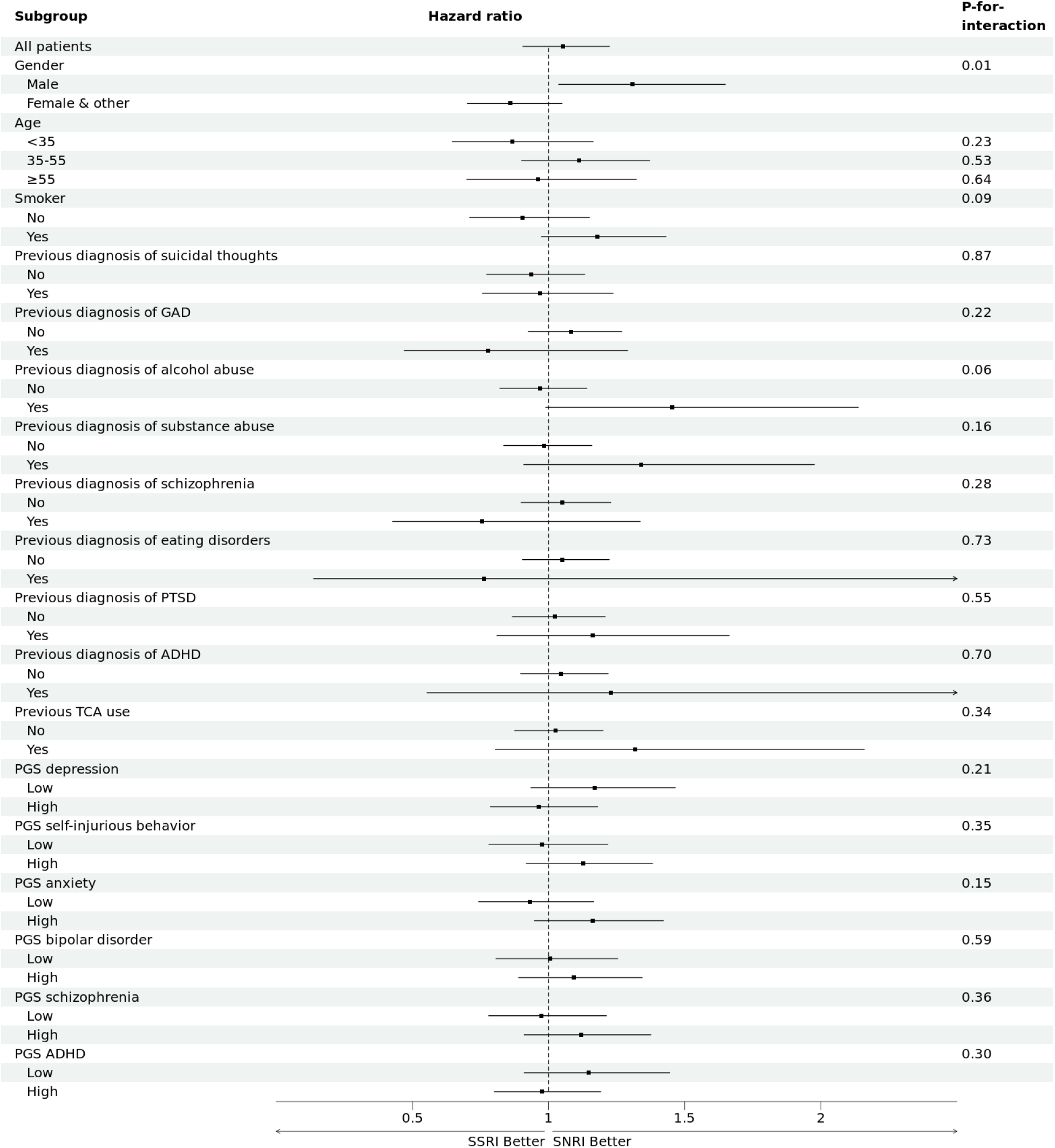
Subgroup analyses for SSRIs vs. SNRIs.

## Discussion

We find evidence that bupropion may be less likely to elicit suicidal thoughts compared to SSRIs among individuals with high genetic predisposition for psychiatric disorders, particularly for ADHD. Notably, conventional subgroup analyses across nongenetic characteristics did not reveal heterogeneity in the risk of suicidal thoughts. These findings suggest that considering PGSs in the personalized medicine framework may help guide clinicians’ choice of antidepressants to avoid serious adverse effects such as suicidal thoughts.

Our finding that bupropion may be less likely to elicit suicidal thoughts among individuals with high ADHD PGS may be related to the fact that bupropion improves ADHD symptoms [19], which includes impulsivity and hyperactivity, and supports the role of impulsivity in suicidal ideation and behavior [20]. Interestingly, we did not find that those who have received a previous diagnosis of ADHD had a lower risk of suicidality with bupropion use compared to those without an ADHD diagnosis. Thus, even if an individual has not clinically manifested ADHD symptoms, merely their genetic tendency to do so may guide clinical practice.

We did not find large heterogeneity in the effects of SSRIs relative to SNRIs across PGSs. Instead, we found that males may have lower risk of increased suicidal thoughts with SNRI (relative to SSRI) use compared to other genders. This is generally in agreement with previous RCTs, which suggest that females may respond better to antidepressants than males and that this trend is more evident for SSRIs than SNRIs [21], though direct comparisons between SSRIs and SNRIs are not well-documented. Thus, this gender difference may be a novel finding that warrants further investigation.

Thus far, it was unclear whether genetic information could guide personalized care for patients with depression. Though there have been associational studies on PGSs for various psychiatric diseases, decisionmaking in psychiatric care was, for the most part, based primarily on patients’ clinical and sociodemographic characteristics and not on their genetic information. Furthermore, even when genetic information is available, few large scale databases have both clinical and genetic data that can evaluate its utility in predicting the adverse events of antidepressants. Combining clinical and genetic information on a large sample of antidepressant users from the All of Us Research Program, the present study filled this knowledge gap on the utility of genetic information in personalized psychiatric care: at least for minimizing suicidal thoughts, PGSs are likely useful in personalizing pharmacotherapy for depression. Investigating this further may provide a better picture of the pathophysiology and subtyping of depression and comorbidities, with which we can further advance personalized psychiatry.

Our study should be interpreted in light of several limitations. First, the PGSs had limited performance in predicting each psychiatric disorder, especially for minoritized populations (though they were comparable to their performance described in their respective original studies). Although this is beyond the scope of the exploratory nature of our study (in addition to the fact that the development of PGSs was not an objective of this study), attempts to improve PGS performance among the general and minoritized populations, such as the one by Lennon and others [22], may be warranted. Second, there may have been misclassification, especially for self-reported variables such as education level, annual income, health insurance coverage, birthplace, and smoking status. Third, participants in the All of Us Research Program may not be generalizable to the general population, as the program obtains data only from patients in registered facilities. Although this may currently be challenging, our findings should be validated in other cohorts in the future. Fourth, as indicated by the wide confidence intervals of the hazard ratios, some estimates of treatment effects are imprecise. As such, the effect heterogeneity elucidated in this study alone is not sufficient to change clinical practice, and more rigorous study designs (such as randomized trials) should be considered to validate our findings. Finally, there may have been unmeasured confounding by individuals’ clinical, sociodemographic, or molecular profiles not evaluated in the All of Us Research Program.

In conclusion, among individuals with major depressive disorder, we detect substantial heterogeneity in the risk of suicidal thoughts with antidepressant use across PGSs for psychiatric disorders. Considering PGSs for various psychiatric disorders may help clinicians tailor antidepressants to each patient to avoid serious adverse effects such as suicidal thoughts.

## Methods

### Study design and data

We aimed to emulate a randomized controlled trial (RCT) to evaluate the effect of antidepressants on suicidal thoughts among patients with major depressive disorder, as well as the heterogeneity in the effects across PGSs for several psychiatric disorders and clinical characteristics of patients. To emulate this hypothetical RCT, we used data from the All of Us Research Program, a large National Institutes of Health-led longitudinal cohort of a diverse sample of the US population [18].

The All of Us Research Program combines participant-derived information from surveys, electronic health records, and biospecimens (including whole genome sequences of blood samples), among other sources, and launched its national recruitment in May 2018. As of 2024, more than 835,000 participants have been enrolled, of whom more than 588,000 had provided biospecimens. Short-read whole genome sequences were available for more than 245,000 of these participants. Enrollment centers are from various settings, including health provider organizations and community partners, with emphasis on recruiting participants from groups historically underrepresented in biomedical research.

All analyses were conducted on the All of Us Researcher Workbench, a cloud-based platform that allows researchers to perform data extraction, curation, and analyses. No individual-level data were downloaded onto a local computer.

### Target trial emulation

We used the target trial emulation framework [17] to emulate a RCT using observational data to obtain treatment effect estimates with minimal bias. We used an active-comparator design for drug effect comparison [23] to align the timings of eligibility evaluation, intervention, and the initiation of the follow-up period. The follow-up period started at treatment assignment and ended at the occurence of the outcome or at five years from treatment assignment, whichever occurred first. All individuals with a diagnosis of major depressive disorder and with available outcome, treatment, and covariate data, including genomic data, available were eligible for matching.

Each participant in the treatment group was matched without replacement to the control group at a 1-to-1 ratio. We used propensity scores to match the participants, with a caliper of 0.10. Propensity scores were calculated by constructing logistic regression models of the treatment using all covariates, and the matched treatment and control samples were compared by calculating the standardized mean differences (SMDs) for each covariate. We included a square term of age in the propensity score model to obtain adequate balance between the covariates. Data on all participants meeting the inclusion criteria were used to construct the models, meaning some participants in the propensity score models were not included in the final matched sample. Matching was performed sequentially in chronological order of the month of antidepressant initiation and was done separately for each month. The order of matching of participants with the same month of antidepressant initiation was chosen at random.

### Variable extraction

Data for all variables were extracted from the All of Us Researcher Workbench. The All of Us Research Program records different diseases and drugs based on the codes in each participating hospital. The codes are thus not uniform: one patient may have a disease coded in International Classification of Disease (ICD)-10, while another may have a disease coded in ICD-9. Thus, the All of Us Research Program uses the Systematized Nomenclature of Medicine (SNOMED) [24] to match up these disease codes. Similarly, the All of Us Research Program records drugs using RxNorm [25], which codes all medications available on the US market. We manually surveyed all SNOMED and RxNorm keywords related to the diseases and drugs of interest in the All of Us Research Program, and chose relevant SNOMED and RxNorm keywords for each disease or drug, as well as the dates they were recorded. The keywords we used are available as a Supplementary Note. All other variables, including sociodemographic variables, were recorded via a survey by the All of Us Research Program.

This study sought to emulate a RCT that performs comparisons of two pairs of antidepressants: SSRIs vs. bupropion, and SSRIs vs. SNRIs. In both comparisons, treatment effects were estimated with SSRIs as the treatment and the other drug as the control.

Participants were allocated into the treatment or control group based on whichever drug was initiated first. For instance, if an individual was started on SSRIs and was later given SNRIs, the individual was classified into the SSRI group. Thus, all analyses of treatment effects were intention-to-treat analyses. If an individual was given both drugs on the same date, they were excluded from the analyses.

The outcome of interest was suicidal thoughts, as recorded in electronic health records. As suicidal ideation is most common with the initiation of antidepressants, we did not evaluate the long-term effects of antidepressants, and the outcome was censored at five years.

We included the following covariates obtained from self-reported surveys in the matching process and evaluation of effect heterogeneity: gender (female, male, or other), age, race and ethnicity (Non-Hispanic Black, Non-Hispanic White, Hispanic, or other), education level (Whether they attended college), annual income (less than $25,000, $25,000 to $50,000, $50,000 to $100,000, or more than $100,000), health insurance coverage, birthplace (US or other), and smoking status (at least 100 cigarettes in one’s lifetime or not). All survey variables were recorded at the time of recruitment into the All of Us Research Program, regardless of the timing of drug initiation.

We also included the previous diagnoses of suicidal thoughts, generalized anxiety disorder, alcohol abuse, schizophrenia, eating disorders, post-traumatic stress disorder (PTSD), and ADHD, as well as the previous use of tricyclic antidepressants. Here, a previous diagnosis or previous use of drug was defined as the coding of the disease or drug in the electronic health record any time before the initiation of the drug of interest. We did not include the previous diagnosis of any other mood disorders.

### Polygenic scores

We obtained short-read whole genome sequences from each All of Us Research Program participant. The All of Us Genome Centers used the same same protocol for library construction (PCR Free Kapa HyperPrep), sequencing (NovaSeq 6000), and software (DRAGEN v3.7.8). The All of Us Research Program applies extensive sample-level and variant-level quality control, including contamination checks, coverage, concordance with genotyping arrays, joint callset quality control, and allele balance filtering. Full details of the sequencing and quality control procedures are described in the All of Us Genomic Quality Report (CDRv8).

For this study, we used the ACAF threshold short-read whole genome sequence callset, a curated subset that contains variants with allele frequency greater than 1% or allele count greater than 100 in at least one ancestry group, mapped to GRCh38. Aside from restricting the callset to individuals in our cohort and applying the PGS scoring described below, no additional variant filtering or imputation was performed.

We included PGSs for depression, self-injurious behavior, bipolar disorder, schizophrenia, anxiety, and attention deficit-hyperactivity disorder (ADHD) as covariates. These were chosen based on previous literature on potential associations between PGSs and treatment effect heterogeneity of antidepressants and psychiatric disorders that have genetic overlap with depression [10, 12, 14, 15], as well as the availability of each PGS on PGS catalog. Of note, the PGS for anxiety included not only generalized anxiety disorder but also panic disorder (episodic paroxysmal anxiety), mixed anxiety and depressive disorder, other mixed anxiety disorders, other specified anxiety disorders, and anxiety disorder, unspecified.

For each trait, we calculated the participants’ PGS using scoring files from PGS Catalog (PGS IDs are provided in Supplementary Table 1), which contain harmonized GRCh38 genomic positions and per-allele effect weights determined by the PGS developers. PGSs were computed as the weighted sum of effect-allele dosages. All scoring was performed using Hail (v0.2.130). Each PGS was standardized across the All of Us Research Program participants with non-missing race and ethnicity and trait labels prior to analyses. For each PGS, we calculated the Nagelkerke’s R-squared value and area under the ROC (AUROC), for the full sample and stratified by race and ethnicity. Evaluation of PGSs were performed using all available participant data; i.e., all participants with available PGSs and outcomes, even if they did not meet the inclusion criteria for the evaluation of treatment effects, were included.

### Statistical analysis

We first compared the risks of suicidal thoughts between SSRI users and bupropion users and between SSRI users and SNRI users by plotting a Kaplan-Meier curve. In addition, we fit Cox proportional hazard models to estimate hazard ratios and 95% confidence intervals (CIs) for the outcome events.

Then, we conducted subgroup analyses to evaluate the heterogeneity in the risks of suicidal thoughts associated with antidepressant use across demographic and clinical characteristics as well as PGSs. Gender was dichotomized as the number of individuals reporting to be neither male nor female were small. Age was trichotomized to group the participants into roughly same subsamples and to capture the possibility that antidepressants respond differently among young populations. PGSs were dichotomized at the median for interpretability. P-values were calculated for the coefficients of the interaction terms between the treatment and covariates of interest.

### Ethical considerations

The All of Us Research Program has been approved by the All of Us Institutional Review Board. As the Program follows a “passport model” that grants broad access to the research dataset approved by the program institutional review board instead of the institutional review board at researchers’ affiliations, the present study was exempt from obtaining ethical approval from authors’ institutions. All researchers who access the All of Us Research Program data are authorized and approved via a process that includes registration, affiliation with an institution that has completed a Data Use and Registration Agreement, identity verification, completion of ethics training, and attestation to a data use agreement. Results reported comply with the All of Us Data and Statistics Dissemination Policy that prohibits disclosure of group counts under 20 to protect participant privacy.

## Data Availability

All data used in this study are available through the All of Us Research Program.

## Acknowledgements

We gratefully acknowledge All of Us participants for their contributions, without whom this research would not have been possible. We also thank the National Institutes of Health’s All of Us Research Program for making available the participant data and/or samples examined in this study.

## Data and code availability

As per All of Us Research Program Data Use Policies, individuals are prohibited from removing participant-level data from the Researcher Workbench. Data analysis code that does not contain participant-level data are available through https://github.com/ryunosukegoto/PGS-HTE.

## Extended Data Figures

**Extended Data Figure 1:**
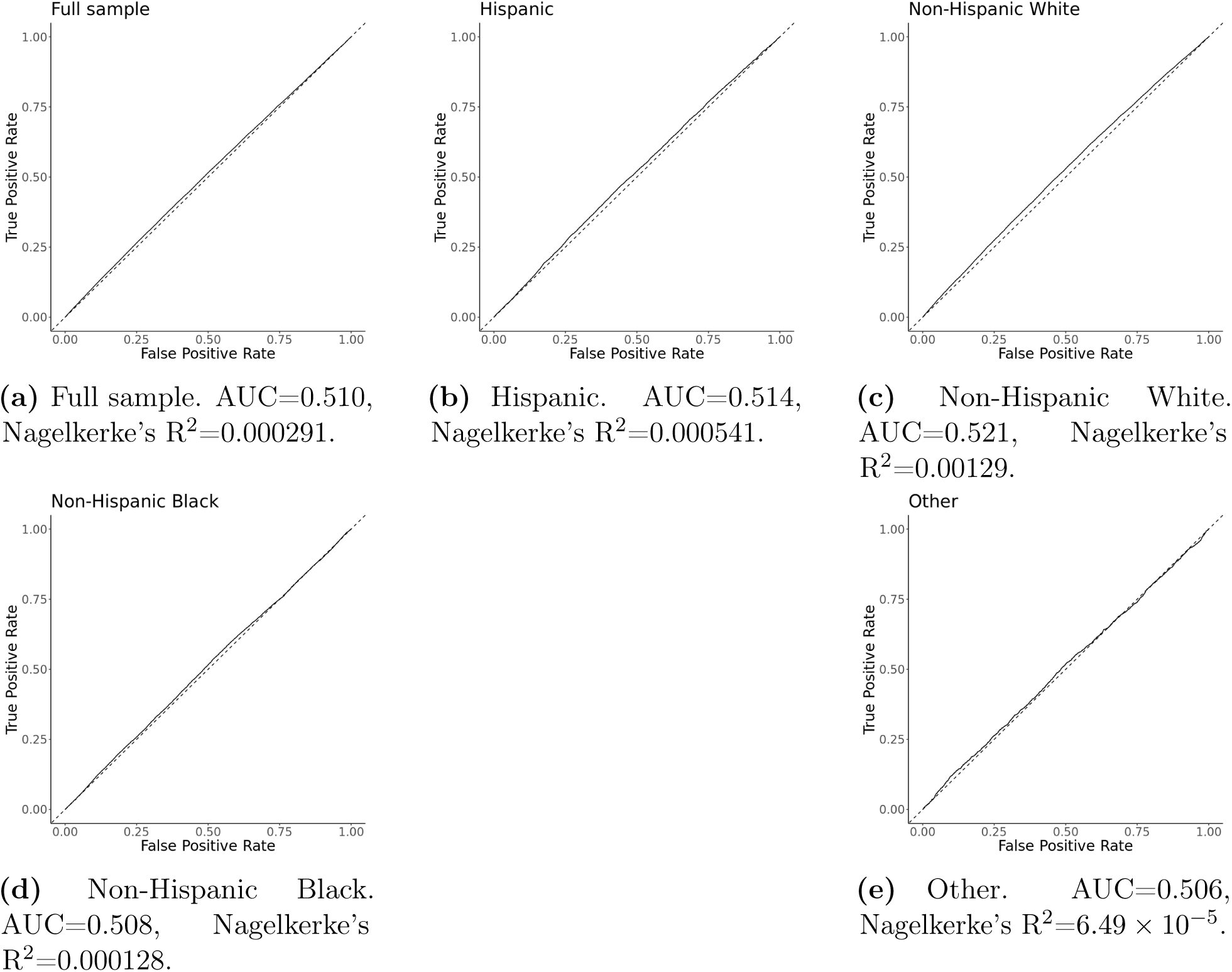
ROC curves of PGS for depression.

**Extended Data Figure 2:**
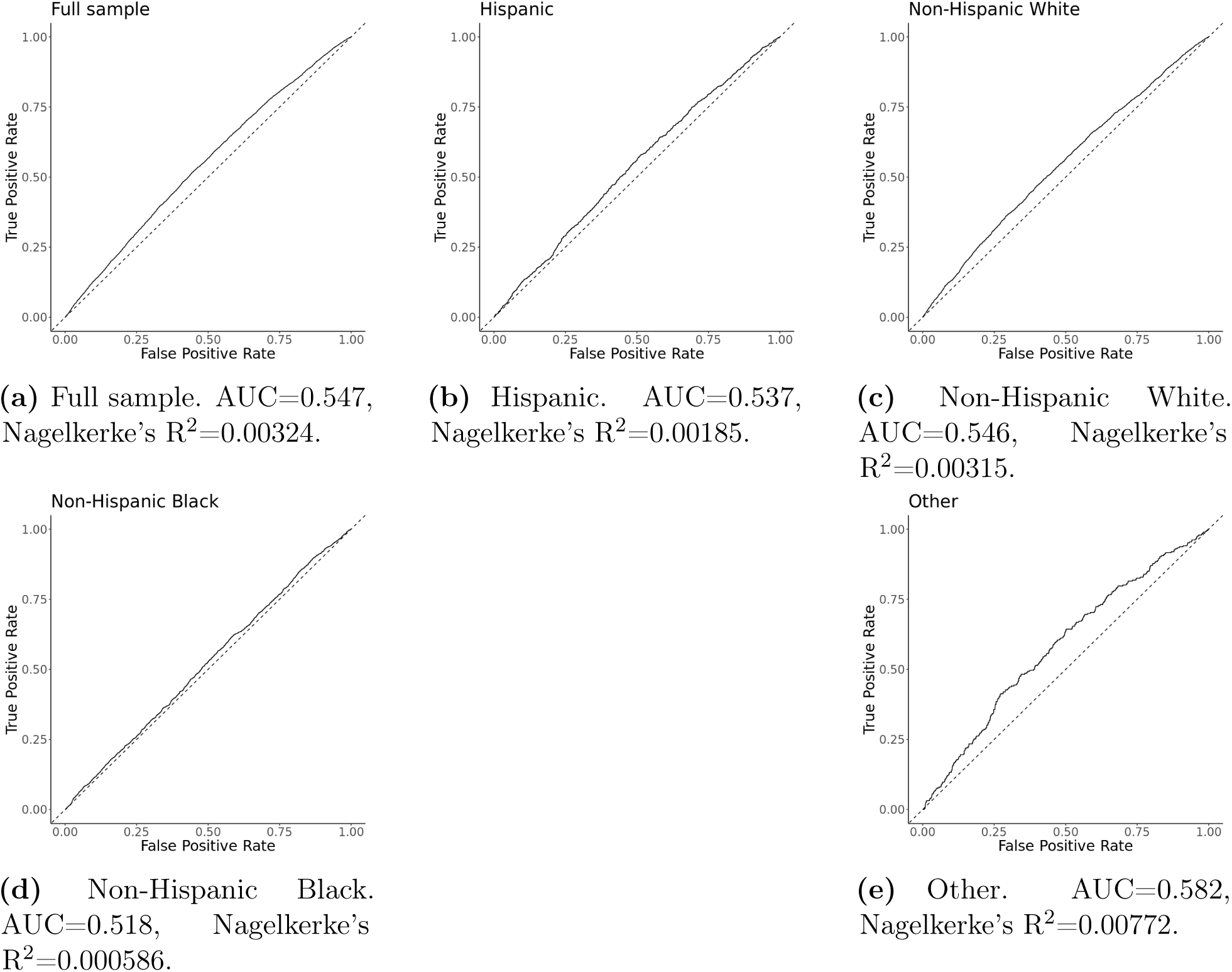
ROC curves of PGS for self-injurious behavior.

**Extended Data Figure 3:**
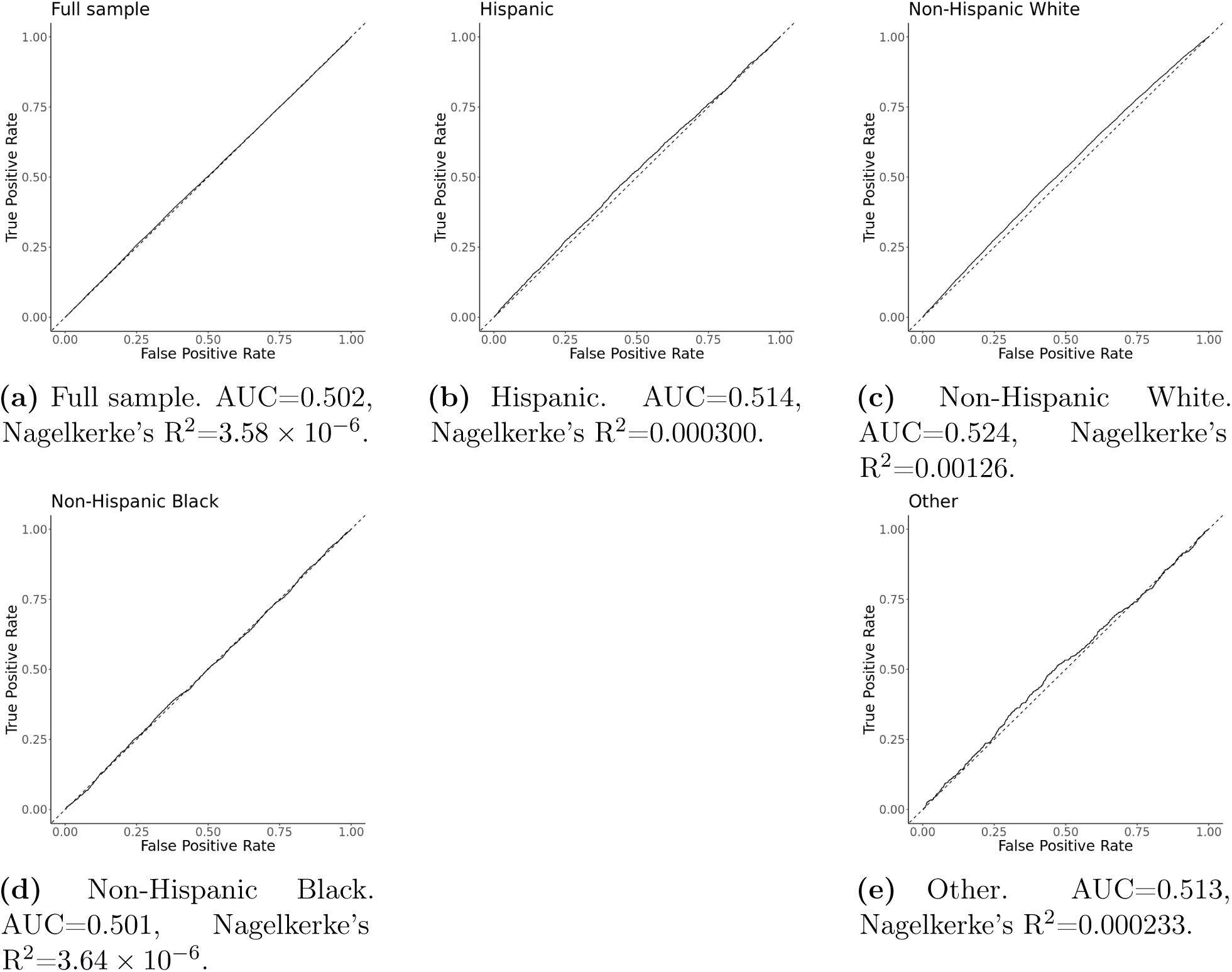
ROC curves of PGS for anxiety.

**Extended Data Figure 4:**
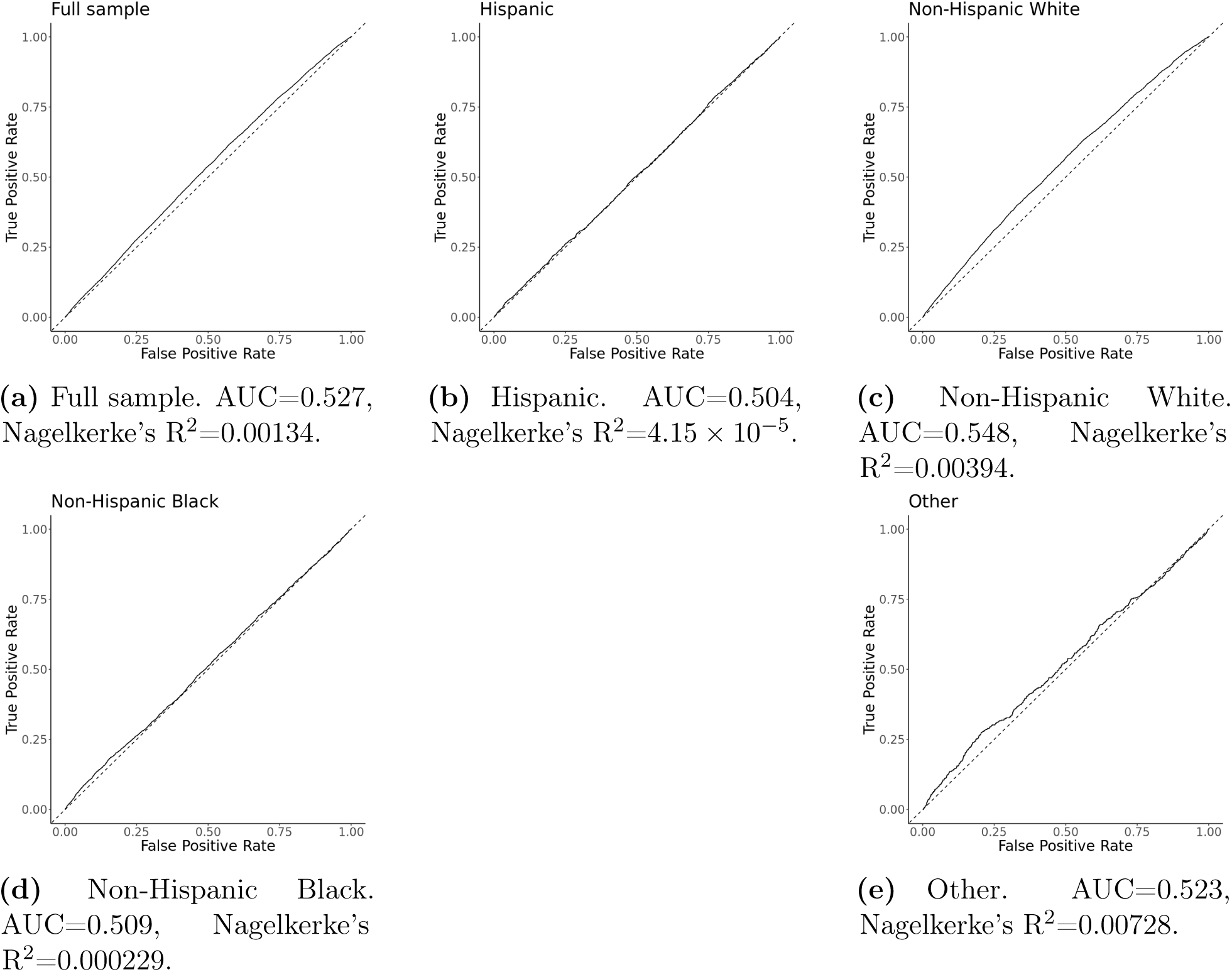
ROC curves of PGS for bipolar disorder.

**Extended Data Figure 5:**
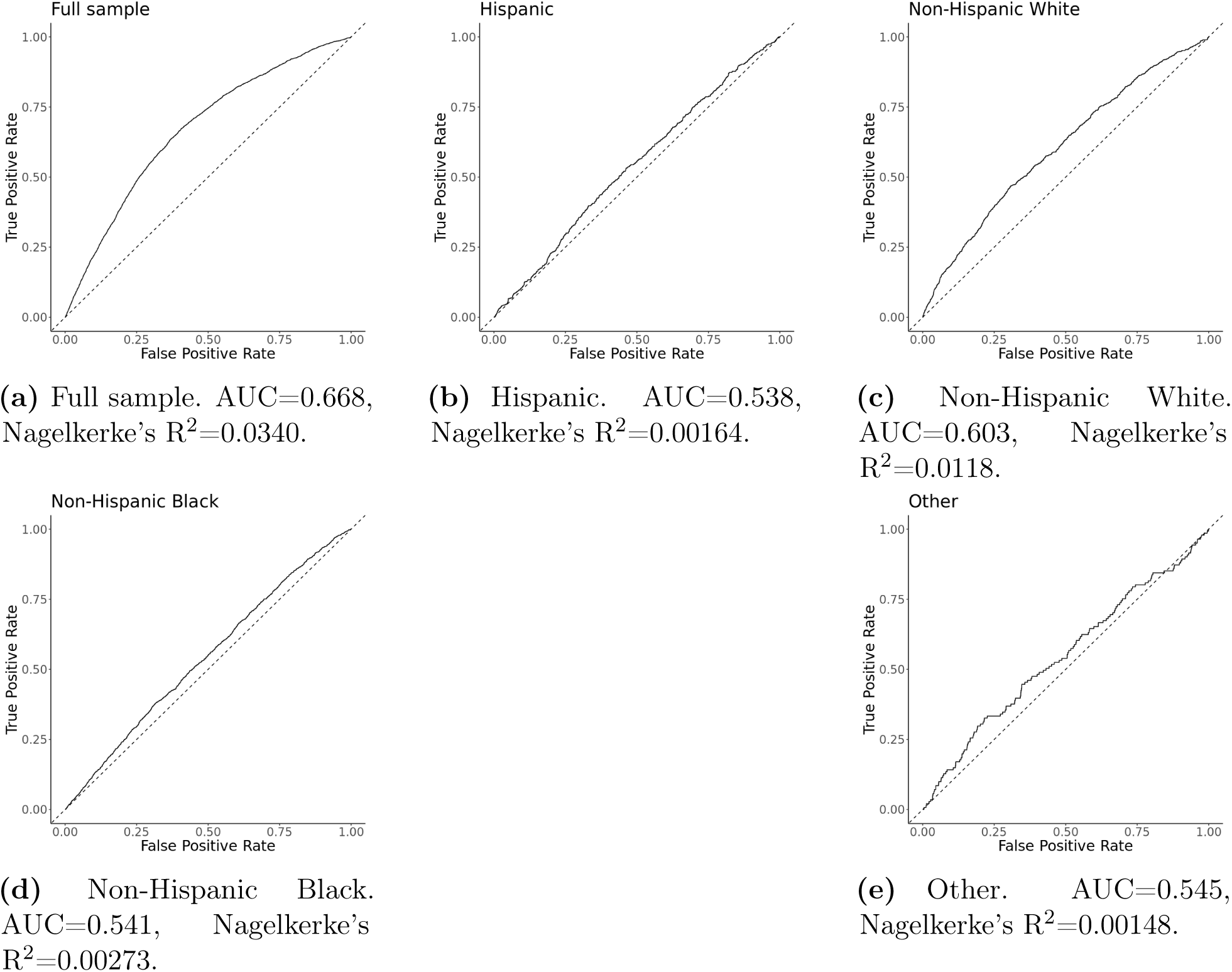
ROC curves of PGS for schizophrenia.

**Extended Data Figure 6:**
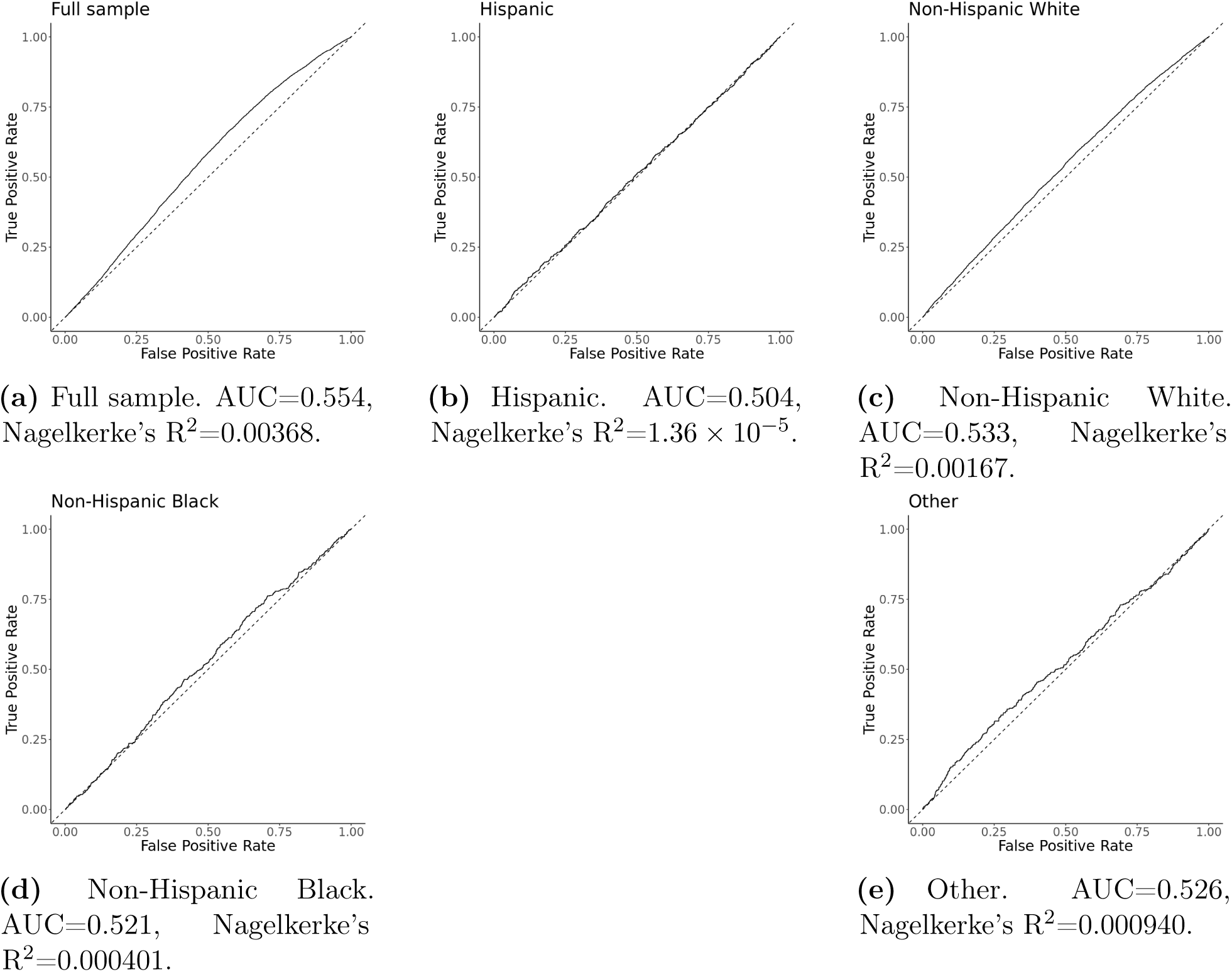
ROC curves of PGS for ADHD.

## Supplementary Note

Keywords for extracting disease and drug information

We used the following keywords to extract disease information:

- Major depressive disorder: contains “major depressive disorder” or “major depression”
- Suicidal thoughts: contains “thoughts of self harm” or “suicidal thoughts” or “planning suicide” or suicidal intent” or “thoughts of deliberate self harm”
- Generalized anxiety disorder: contains “generalized anxiety disorder”
- Bipolar disorder: contains “bipolar”
- Alcohol abuse: contains “alcohol abuse”
- Substance abuse: contains “amphetamine abuse” or “cocaine abuse” or “cannabis abuse”
- Schizophrenia: contains “schizophrenia”
- Eating disorders: contains “anorexia nervosa” or “bulimia nervosa”
- Post-traumatic stress disorder: contains “posttraumatic stress disorder” or “post-traumatic stress disorder”
- Attention deficit-hyperactivity disorder: contains “attention deficit hyperactivity disorder”

We used the following keywords to extract drug information:

Selective serotonin reuptake inhibitors: contains “citalopram” or “escitalopram” or “fluoxetine” or “fluvoxamine” or “paroxetine” or “sertraline”
Serotonin and norepinephrine reuptake inhibitors: contains “desvenlafaxine” or “duloxetine” or “milnacipran” or “venlafaxine”
Bupropion: contains “bupropion”
Tricyclic antidepressants: contains “amitriptyline” or “amoxapine” or “clomipramine” or “desipramine” or “doxepin” or “imipramine” or “maprotiline” or “nortriptyline” or “protriptyline” or “trimipramine”

## Supplementary Tables

**Supplementary Table 1:** Traits, polygenic score ID, and URL of the polygenic scores.

| Trait | PGS ID | URL |
| --- | --- | --- |
| Depression | PGS000145 | https://www.pgscatalog.org/trait/MONDO_0002009/ |
| Self-injurious behavior | PGS002222 | https://www.pgscatalog.org/trait/HP_0100716/ |
| Anxiety<sup>c</sup> | PGS004451 | https://www.pgscatalog.org/trait/EFO_0006788/ |
| Bipolar disorder | PGS002786 | https://www.pgscatalog.org/trait/MONDO_0004985/ |
| Schizophrenia | PGS002785 | https://www.pgscatalog.org/trait/MONDO_0005090/ |
| ADHD | PGS002746 | https://www.pgscatalog.org/trait/EFO_0003888/ |

<sup>c</sup> Includes panic disorder (episodic paroxysmal anxiety), generalized anxiety disorder, mixed anxiety and depressive disorder, other mixed anxiety disorders, other specified anxiety disorders, and anxiety disorder, unspecified. PGS, polygenic score; ADHD, attention deficit-hyperactivity disorder.

